# COVID-19 collateral damage: psychological distress and behavioral changes among older adults during the first outbreak in Stockholm, Sweden

**DOI:** 10.1101/2021.03.16.21253750

**Authors:** Giorgi Beridze, Federico Triolo, Giulia Grande, Laura Fratiglioni, Amaia Calderón-Larrañaga

## Abstract

**Introduction:** During the first wave of the COVID-19 pandemic, Swedish public health authorities provided recommendations for 70+ year old people. They were strongly encouraged to self-isolate but remain physically active in a safe manner. This study aimed to explore the indirect, negative effects of COVID-19 restrictions (collateral damage) by exploring to what extent adherence to such recommendations might have impacted the lives and health of older adults living in central Stockholm.

**Methods:** An *ad-hoc* phone questionnaire was administered by trained staff between May and June 2020 to a random sample of older adults 68+ years old (n=1231), who had attended the regular follow-up assessment of the longitudinal Swedish National study on Aging and Care in Kungsholmen (SNAC-K) during 2016-2019. We explored three dimensions of collateral damage, namely psychological distress (feelings of worry, stress and loneliness), reductions in social and physical activities, and reductions in medical and social care use. Logistic regression models were used to test the association between age, sex, education and living arrangement, and the risk of collateral damage.

**Results:** Vast majority of participants adhered to the recommendations, with over three quarters practicing self-isolation (n=928). Half of the sample reported psychological distress, 55.3% reported reductions in social or physical activity, and 11.3% reported decreased medical or social care use. Over three quarters of participants were affected by at least one of the three collateral damage dimensions. Female sex was the strongest sociodemographic predictor of individual as well as co-occurring dimensions of collateral damage.

**Conclusion:** COVID-19 and its restrictions during the first half of 2020 have had a negative effect on the health and lives of a majority of elderly living in central Stockholm. Women were at a particularly higher risk of these negative consequences. We emphasize the need for predefined, evidence-based interventions to address these negative consequences.

**STRENGTHS AND LIMITATIONS OF THIS STUDY:**

- This study uses a large sample of older adults from a well-characterized population-based study (SNAC-K)
- Several dimensions of the indirect, negative effects (collateral damage) of the COVID-19 pandemic are explored in this study
- As Sweden’s response to COVID-19 differed from most countries, this study provides a unique opportunity of comparison with other settings
- The cross-sectional design of this study does not allow to establish temporality between the onset of the pandemic and studied outcomes
- The results of this study may not be generalisable to the entire elderly population in Sweden as participants are from an urban, affluent area in Stockholm

## INTRODUCTION

The outbreak caused by the novel coronavirus, SARS-CoV2, was declared a global pandemic by the World Health Organization (WHO) on March 11^th^, 2020, coinciding with the date of the first confirmed death in Sweden. Early on, it was identified that older adults are at a significantly higher risk of mortality from COVID-19. Indeed, as of January 18^th^ 2021, 91% of deaths attributed to COVID-19 in Sweden have happened in those aged 70 and above [1]. Later on, additional prognostic factors were identified, including male sex, socioeconomic disadvantage, the presence of comorbidities and frailty [2–5].

In response to the pandemic, most countries have implemented strict measures to help curb the spread of the virus and reduce mortality. Sweden’s response to COVID-19 differed from most countries by not implementing strict lockdowns and restrictions, but instead relying on high voluntary adherence to the recommendations proposed by the Public Health Agency. The general recommendations included keeping good hand hygiene, practicing social distancing and avoiding contact if having any symptoms [6]. On top of these, the specific recommendations for the elderly were to stay at home, avoid social gatherings and public transportation, but to remain physically active outdoors in a safe manner [6].

The importance of looking beyond mortality and morbidity when assessing national response strategies for COVID-19 has been raised [7]. Stay-at-home orders and lack of contact with loved ones put elderly at risk of loneliness and social isolation, which, in turn, are known to have negative effects, particularly in old age [8]. Reduced physical activity and sedentarism have detrimental effects on physical and mental health [9,10]. Additionally, due to the overburdening of healthcare services and reduced access to medical, social and informal care, new conditions may not be timely diagnosed, and existing health conditions may be exacerbated. These consequences can be considered indirect, negative effects (i.e. collateral damage) of COVID-19 restrictions.

While several studies have examined the distribution of COVID-19 mortality among Swedish older adults by sex, socioeconomic and household factors [11–13], little is known on the collateral damage of these restrictions. To the best of our knowledge, only one study has examined the mental health consequences of the Swedish strategy on the elderly [14], and no study has looked into other dimensions such as psychological wellbeing and/or behavioral changes. Thus, the aims of this study are to explore different dimensions of the collateral damage linked to COVID-19 during the first epidemic outbreak in an older population of central Stockholm, as well as to characterize the sociodemographic profile of those with the highest susceptibility to this damage.

## METHODS

### Study population

Study population consisted of 1231 older adults aged between 68 and 103, participating in the Swedish National study on Aging and Care in Kungsholmen (SNAC-K). SNAC-K is a longitudinal study including a random sample of older adults aged 60 years and above living in the Kungsholmen area of Stockholm, Sweden. All SNAC-K participants who had participated in the regular follow-up assessment during 2016-2019 were invited to participate in a telephone interview aimed at monitoring preventive behaviors and the direct and indirect health consequences of the COVID-19 pandemic. Telephone interviews were conducted between May and June (95%) by trained SNAC-K staff, following a multi-choice questionnaire that was *ad hoc* developed by SNAC-K researchers. Exclusion criteria included living in care and nursing homes, known diagnosis of dementia and very impaired hearing. The response rate was 91.9%. Subjects who refused to participate or could not be contacted had similar age and educational attainment to those who participated, but were more likely to be male (45.4% vs 35.7%, p=0.044).

### Collateral damage

In this study we examined three dimensions of collateral damage: psychological distress and two aspects related to behavioral changes, i.e. reductions in social and physical activities, and in medical and social care use. Psychological distress was assessed with variables related to worrying about being affected by COVID-19 (very/extremely vs. not at all/somewhat/moderately), worrying about loved ones being affected by COVID-19 (very/extremely vs. not at all/somewhat/moderately), feeling nervous and/or stressed (often/very often vs. never/sometimes), and loneliness (≥5 vs. <5 on the Three-Item Loneliness Scale [15]). Changes in social and physical activities were measured by asking participants about reductions in social interactions, reductions in light physical activity (yes/no) and reductions in vigorous physical activity (yes/no). A reduction in social interactions, hereon referred to as social isolation, was defined as a reduction in physical communication with family and friends without an increase in phone and/or video communication. Care-related items included refraining from seeking medical care (yes/no) and receiving reduced care at home. Reduced care at home was defined as a decrease in the use of formal home-care services without an increase in received informal care.

### Preventive measures and sources of information

Participants were asked about their adherence to a list of 9 recommendations −both general and those specific to elderly−, and the most common sources of information regarding the COVID-19 pandemic.

### Sociodemographics

Sociodemographic variables in the present study included age, sex, education and living arrangement. Age was dichotomized as youngest old (≤80 years old) and oldest old (>80 years old). Highest obtained education was dichotomized as low (elementary) and high (high school, university, or above). Living arrangement was dichotomized into those who lived alone and those who did not.

### Statistical analysis

Characteristics of the study sample were reported as overall, as well as stratified by the four sociodemographic variables. Between-group differences were assessed via two-tailed t-tests and chi-square tests as appropriate. Binary logistic regression models were used to assess the associations between sociodemographic variables and the different collateral damage dimensions, as well as each item within these dimensions. All models were mutually adjusted for all sociodemographic variables. All statistical tests were performed in StataSE 15 (StataCorp LLC, College Station, TX, USA). Significance level (alpha) was set at 0.05 for all analyses.

### Ethical considerations

Informed consent was obtained from all participants. The study was approved by the Regional Ethics Review Board in Stockholm (dnr: 2020-02497).

### Patient and public involvement

There was no direct public involvement either in the setting of the research questions or developing the study design.

## RESULTS

The mean age of participants was 78.2 years, 64.3% were female, 34.3% had elementary educational attainment and 50.2% lived alone (**Table 1**). Five percent of participants (n=62) reported being tested for COVID-19, 9 of which reported testing positive. Almost half of the sample (48.3%, n=595) sought medical care during the period March-June 2020, with 79 of them finding it more difficult to access it. Nine participants reported being hospitalized for suspected or confirmed COVID-19.

**Table 1.** Study sample characteristics (N=1231).

|  |  |
| --- | --- |
| **Age** mean (SD) | 78.2 (8.3) |
| **Age** n (%) |  |
| ≤80 years | 642 (52.2%) |
| >80 years | 589 (47.8%) |
| **Female** n (%) | 792 (64.3%) |
| **Education** n (%) |  |
| High school, university or above | 809 (65.7%) |
| Elementary school | 422 (34.3%) |
| **Living alone** n (%) | 616 (50.2%) |
| **COVID-19-related symptoms** n (%) |  |
| 0 | 801 (65.1%) |
| 1 | 183 (14.9%) |
| 2+ | 247 (20.0%) |
| **Tested for COVID-19** n (%) |  |
| Yes, positive | 9 (0.7%) |
| Yes, negative/unknown | 53 (4.3%) |
| **Sought medical care** n (%) | 595 (48.6%) |
| **Found it difficult to access healthcare services<sup>a</sup>** n (%) | 79 (14.6%) |
| **Hospitalized due to confirmed or suspected COVID-19** n (%) | 9 (0.7%) |

<sup>a</sup> Subsample of those who sought medical care (n=540)

The most commonly reported preventive behaviors were physical distancing of at least two meters (98.0%) and washing hands for at least 20 seconds (98.0%), followed by covering mouth and nose when coughing or sneezing (88.5%) and staying home in case of illness or cold (88.4%) (**Figure 1**). Three quarters of the sample (76.8%) reported self-isolating. The least commonly reported measure was usage of face masks (15.2%). Most participants stayed up to date on the COVID-19 pandemic using television (95.9%); over three quarters (77.6%) reported following the Public Health Agency’s press-conferences. Digital sources, such as social media and online news websites were the least reported sources (22.9% and 59.8% respectively).

**Figure 1.**
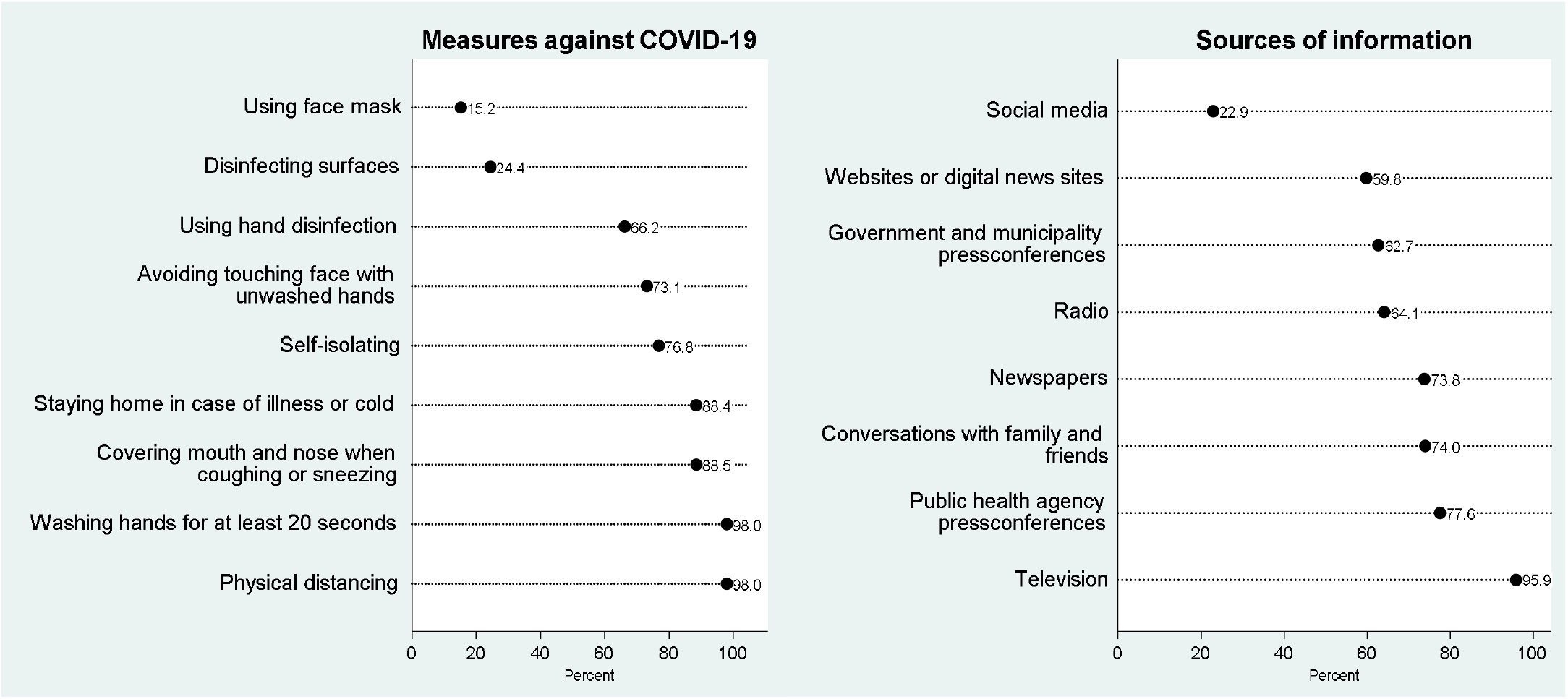
Adherence to preventive behaviors (left) and sources of information (right) related to COVID-19 during the first outbreak in Stockholm (March-June 2020).

Half of the sample experienced psychological distress, with the most common items being loneliness (33.4%) and worrying about loved ones getting COVID-19 (24.9%) (**Table 2**). More than half (55.3%) experienced a reduction in either social or physical activities, and 11.3% either refrained from seeking medical care or received less social care at home. In total, 77.8% of participants (n=956) experienced at least one of the three dimensions of collateral damage comprising psychological distress, reductions in social and physical activities, and decreased medical and social care use. Almost half (43.7%) reported experiencing one dimension of collateral damage, while the remainder (34.1%) experienced two or more. Univariate associations between each of the four sociodemographic variables and the different collateral damage dimensions are presented in **Supplementary Tables 1** and **2**.

**Table 2.**
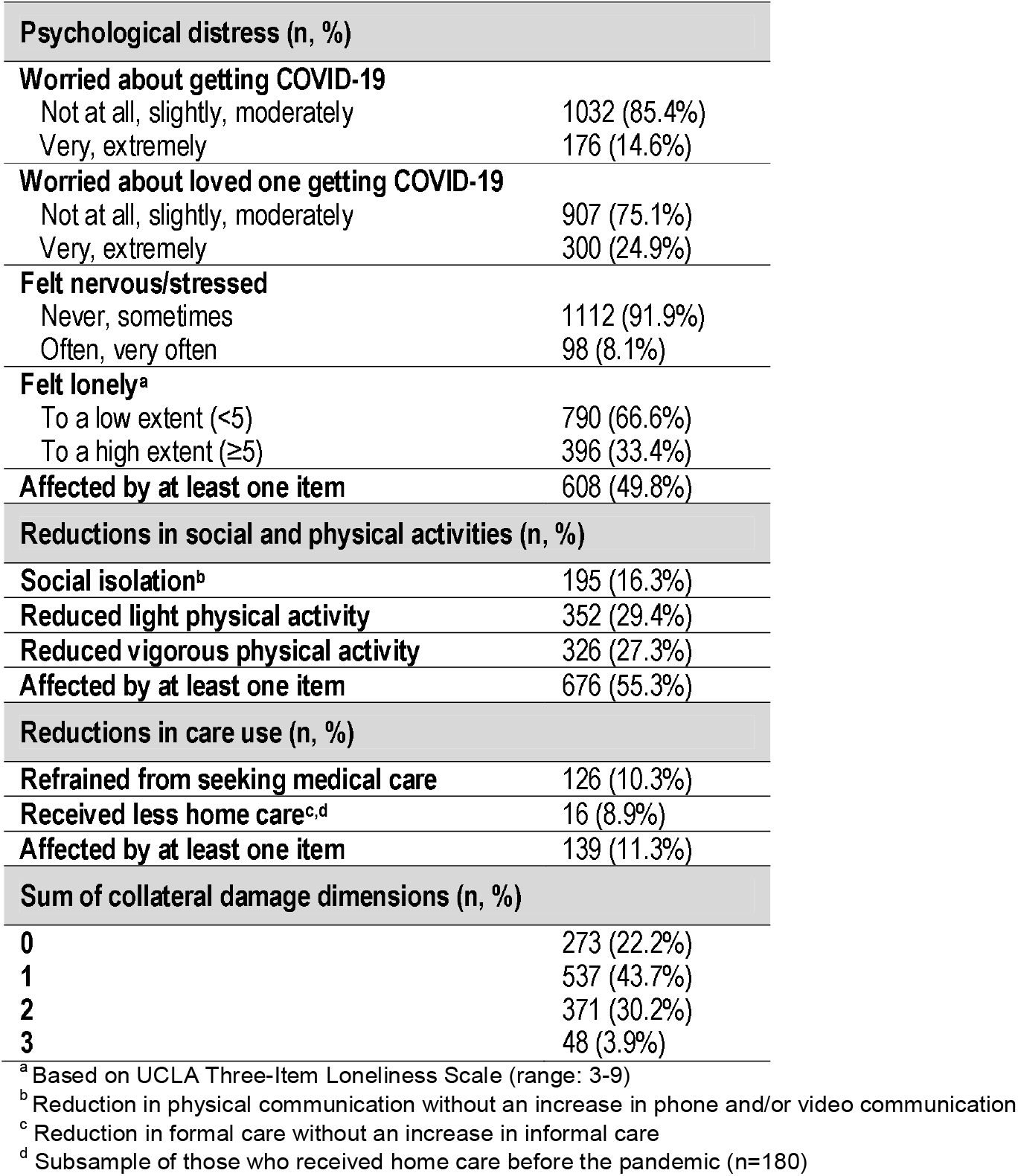
Psychological distress and behavioral changes in the study sample (N=1231) during the first COVID-19 outbreak in Stockholm (March-June 2020).

| Psychological distress (n, %) |  |
| --- | --- |
| **Worried about getting COVID-19** |  |
| Not at all, slightly, moderately | 1032 (85.4%) |
| Very, extremely | 176 (14.6%) |
| **Worried about loved one getting COVID-19** |  |
| Not at all, slightly, moderately | 907 (75.1%) |
| Very, extremely | 300 (24.9%) |
| **Felt nervous/stressed** |  |
| Never, sometimes | 1112 (91.9%) |
| Often, very often | 98 (8.1%) |
| **Felt lonely<sup>a</sup>** |  |
| To a low extent (<5) | 790 (66.6%) |
| To a high extent (≥5) | 396 (33.4%) |
| **Affected by at least one item** | 608 (49.8%) |
| Reductions in social and physical activities (n, %) |  |
| **Social isolation<sup>b</sup>** | 195 (16.3%) |
| **Reduced light physical activity** | 352 (29.4%) |
| **Reduced vigorous physical activity** | 326 (27.3%) |
| **Affected by at least one item** | 676 (55.3%) |
| Reductions in care use (n, %) |  |
| **Refrained from seeking medical care** | 126 (10.3%) |
| **Received less home care<sup>c, d</sup>** | 16 (8.9%) |
| **Affected by at least one item** | 139 (11.3%) |
| Sum of collateral damage dimensions (n, %) |  |
| **0** | 273 (22.2%) |
| **1** | 537 (43.7%) |
| **2** | 371 (30.2%) |
| **3** | 48 (3.9%) |

<sup>a</sup> Based on UCLA Three-Item Loneliness Scale (range: 3-9)
<sup>b</sup> Reduction in physical communication without an increase in phone and/or video communication
<sup>c</sup> Reduction in formal care without an increase in informal care
<sup>d</sup> Subsample of those who received home care before the pandemic (n=180)

Women had higher odds of experiencing all items within the dimensions of psychological distress and social and physical activity reduction (**Table 3**). Within the dimension of psychological distress, the oldest old had significantly lower odds of worrying about getting COVID-19. Those who lived alone had significantly lower odds of worrying about loved ones getting COVID-19, but higher odds of loneliness. Within the dimension of social and physical activity reductions, the oldest old had significantly higher odds of reducing light physical activity, while the oldest old and those who lived alone had lower odds of decreasing vigorous activity. Within the dimension of medical and social care use reduction, those with lower education had higher odds of receiving less care at home.

**Table 3.**
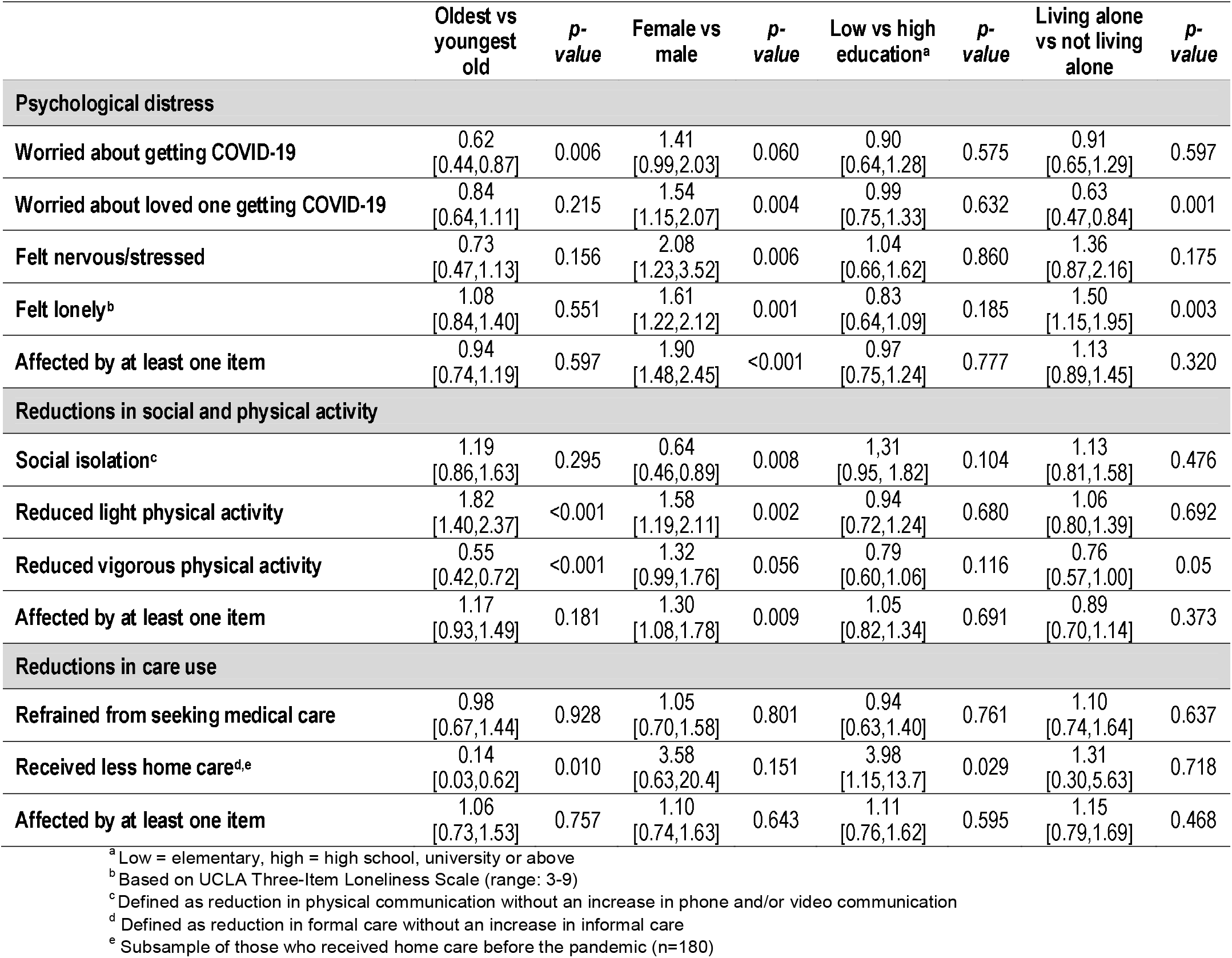
Association (odds ratios and 95% confidence intervals) between psychological distress and behavioral changes and sociodemographic factors (N=1231) during the first COVID-19 outbreak in Stockholm (March-June 2020). Models mutually adjusted by all sociodemographic factors.

|  | Oldest vs youngest old | *p*-value | Female vs male | *p*-value | Low vs high education<sup>a</sup> | *p*-value | Living alone vs not living alone | *p*-value |
| --- | --- | --- | --- | --- | --- | --- | --- | --- |
| **Psychological distress** |  |  |  |  |  |  |  |  |
| Worried about getting COVID-19 | 0.62<br>[0.44,0.87] | 0.006 | 1.41<br>[0.99,2.03] | 0.060 | 0.90<br>[0.64,1.28] | 0.575 | 0.91<br>[0.65,1.29] | 0.597 |
| Worried about loved one getting COVID-19 | 0.84<br>[0.64,1.11] | 0.215 | 1.54<br>[1.15,2.07] | 0.004 | 0.99<br>[0.75,1.33] | 0.632 | 0.63<br>[0.47,0.84] | 0.001 |
| Felt nervous/stressed | 0.73<br>[0.47,1.13] | 0.156 | 2.08<br>[1.23,3.52] | 0.006 | 1.04<br>[0.66,1.62] | 0.860 | 1.36<br>[0.87,2.16] | 0.175 |
| Felt lonely<sup>b</sup> | 1.08<br>[0.84,1.40] | 0.551 | 1.61<br>[1.22,2.12] | 0.001 | 0.83<br>[0.64,1.09] | 0.185 | 1.50<br>[1.15,1.95] | 0.003 |
| Affected by at least one item | 0.94<br>[0.74,1.19] | 0.597 | 1.90<br>[1.48,2.45] | <0.001 | 0.97<br>[0.75,1.24] | 0.777 | 1.13<br>[0.89,1.45] | 0.320 |
| **Reductions in social and physical activity** |  |  |  |  |  |  |  |  |
| Social isolation<sup>c</sup> | 1.19<br>[0.86,1.63] | 0.295 | 0.64<br>[0.46,0.89] | 0.008 | 1.31<br>[0.95,1.82] | 0.104 | 1.13<br>[0.81,1.58] | 0.476 |
| Reduced light physical activity | 1.82<br>[1.40,2.37] | <0.001 | 1.58<br>[1.19,2.11] | 0.002 | 0.94<br>[0.72,1.24] | 0.680 | 1.06<br>[0.80,1.39] | 0.692 |
| Reduced vigorous physical activity | 0.55<br>[0.42,0.72] | <0.001 | 1.32<br>[0.99,1.76] | 0.056 | 0.79<br>[0.60,1.06] | 0.116 | 0.76<br>[0.57,1.00] | 0.05 |
| Affected by at least one item | 1.17<br>[0.93,1.49] | 0.181 | 1.30<br>[1.08,1.78] | 0.009 | 1.05<br>[0.82,1.34] | 0.691 | 0.89<br>[0.70,1.14] | 0.373 |
| **Reductions in care use** |  |  |  |  |  |  |  |  |
| Refrained from seeking medical care | 0.98<br>[0.67,1.44] | 0.928 | 1.05<br>[0.70,1.58] | 0.801 | 0.94<br>[0.63,1.40] | 0.761 | 1.10<br>[0.74,1.64] | 0.637 |
| Received less home care<sup>d,e</sup> | 0.14<br>[0.03,0.62] | 0.010 | 3.58<br>[0.63,20.4] | 0.151 | 3.98<br>[1.15,13.7] | 0.029 | 1.31<br>[0.30,5.63] | 0.718 |
| Affected by at least one item | 1.06<br>[0.73,1.53] | 0.757 | 1.10<br>[0.74,1.63] | 0.643 | 1.11<br>[0.76,1.62] | 0.595 | 1.15<br>[0.79,1.69] | 0.468 |

<sup>a</sup> Low = elementary, high = high school, university or above
<sup>b</sup> Based on UCLA Three-Item Loneliness Scale (range: 3-9)
<sup>c</sup> Defined as reduction in physical communication without an increase in phone and/or video communication
<sup>d</sup> Defined as reduction in formal care without an increase in informal care
<sup>e</sup> Subsample of those who received home care before the pandemic (n=180)

Women were more likely to experience one (OR: 1.38, 95% CI: 1.01;1.90), two (OR: 2.36, 95% CI: 1.66;3.35) and all three (OR: 2.21, 95% CI: 1.08;4.55) collateral damage dimensions compared to men (**Figure 2**). No statistically significant differences were detected for age, education and living arrangement in terms of the number of experienced dimensions of collateral damage, after adjusting for the rest of sociodemographic factors.

**Figure 2.**
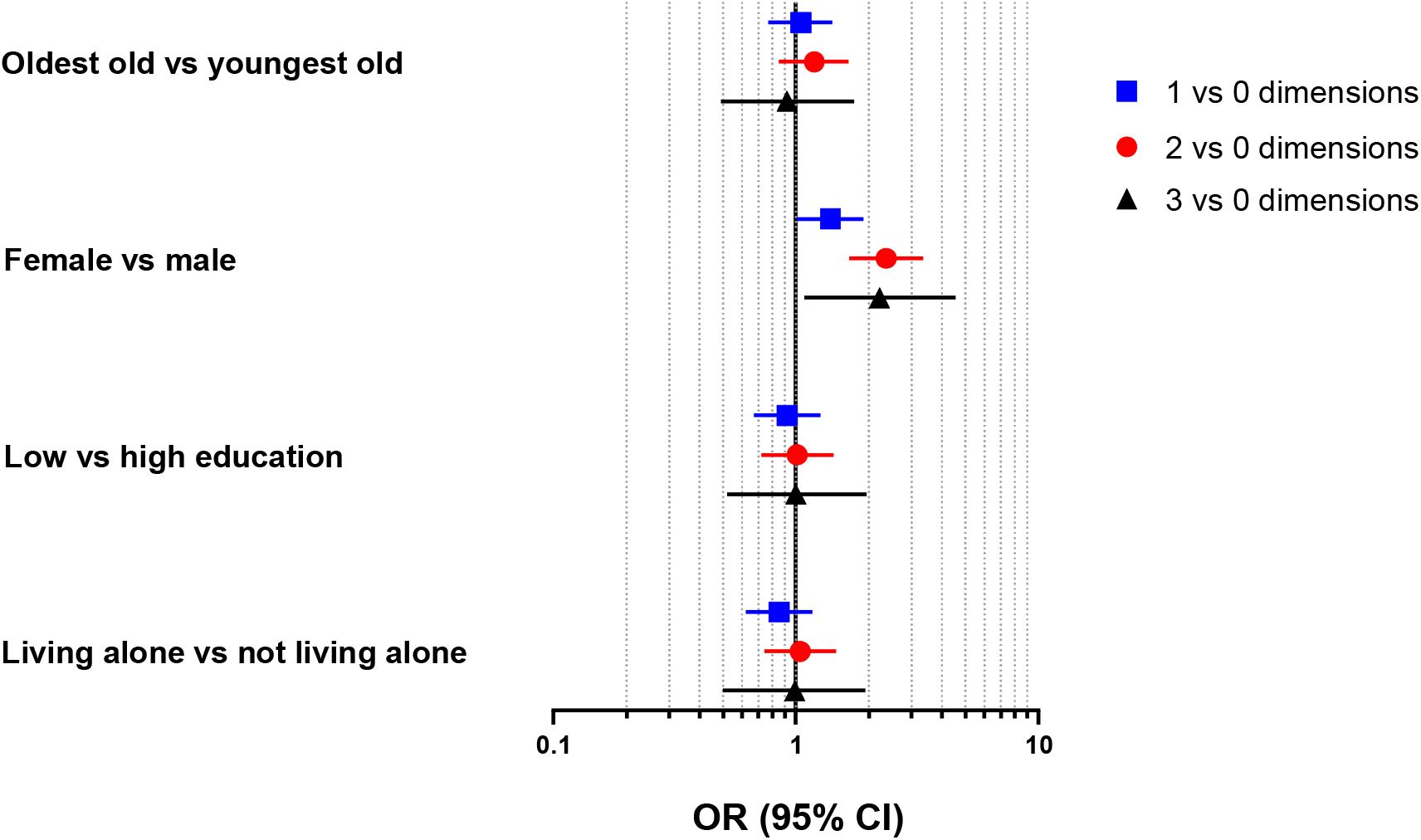
Association (odds ratios and 95% confidence intervals) between number of experienced dimensions of collateral damage and sociodemographic factors (N=1231) during the first COVID-19 outbreak in Stockholm (March-June 2020). Models mutually adjusted by all sociodemographic factors.

## DISCUSSION

This study examined the collateral damage of COVID-19 restrictions on the health and lives of older adults living in Stockholm during the first outbreak. We took into account three dimensions of damage: psychological distress, reductions in social and physical activities, and decreased medial and social care use. We found that over three quarters of the sample was affected by at least one dimension, with women being at a considerably higher risk. The study also provides valuable insights into older people’s adherence to COVID-19 preventive behaviors during the first half of 2020.

### Interpretation of results

Adherence to national recommendations is an important factor in mitigating the negative effects of the COVID-19 pandemic. We found that participants in our study were well-informed about the pandemic and adopted low-risk behaviors during the first wave of COVID-19 outbreak. The majority of participants followed the Public Health Agency press-conferences, likely reflecting the high social and institutional trust in Sweden [14], and adhered to the Agency’s strongly recommended preventive measures.

We observe a substantial impact of the pandemic on the mental health of the elderly, with half of the sample reporting psychological distress. A fair share of the sample reported worrying about themselves and their loved ones being affected by COVID-19. Interestingly, the latter seems to be of more concern, a finding that has been replicated in another Swedish survey [14]. Loneliness and feelings of stress were also prevalent in our sample. This is in line with a large body of research showing considerable effects of the pandemic on mental health outcomes [16,17]. The burden seems to be unevenly borne by women and, to a lesser extent, by those living alone. Indeed, previous research has shown that women are at a higher risk of poor mental health [17] and worrying about family [18] during the pandemic.

High adherence to self-isolation recommendations, combined with a decrease in physical contact with loved ones, puts older adults at risk of social isolation. Social isolation presents a major modern-day challenge and has been associated with several negative health outcomes, such as depression [19], frailty [20], cognitive decline [21] and low quality of life [22,23]. While we did observe a reduction in frequency of physical meetings with family, friends and neighbors, this was largely met with an increase in phone and video communication with them. This is very important in the context of preventing the negative effects of loneliness and social isolation, as alternate forms of communication may buffer such effects [24,25].

Concern has been raised about reduction in physical activity as a major collateral consequence of the pandemic restrictions, as low physical activity is linked to negative cardiovascular and metabolic outcomes [9], poor mental health [10], frailty [26], and insomnia [27], among others. In spite of the Public Health Agency’s recommendations for the elderly to remain physically active and spend time outdoors in a safe manner, we still found that up to a third of the sample had decreased their frequency of both light and vigorous physical activity. Furthermore, it is important to highlight that the reduction in light physical activity was more prominent among the oldest old, who, in all likelihood, were also doing less incidental physical activity, such as climbing the stairs or visiting the supermarket, due to self-isolation. They might encounter difficulties in returning to their former activity levels should the pandemic persist for a long time, which requires close monitoring from medical and social services.

We found that subjects in our sample limited their contact with the healthcare system during the first wave of the pandemic, but when seeking for help, received it in a timely and satisfactory way. This is an important finding in a context where hospital overcrowding has emerged as an important challenge in many countries. Around 10% of our sample refrained from seeking medical care altogether, which may explain, among others, the reduction in cancer diagnosis by Swedish healthcare services compared to previous years [28]. Still, the proportion of those refraining is significantly lower than in the US, where a third of the population aged 65+ reported delaying or avoiding medical care during the first wave of the pandemic [29]. Subjects also decreased their use of formal care but seem to have compensated for it by an increase in received informal care. This could become a concern should the pandemic persist, since it is widely acknowledged that informal caregiving places significant economic, physical and mental burden on caregivers, who are often themselves older adults with health needs [30].

### Strengths and limitations

To the best of our knowledge, this study is the first to examine the consequences of the Swedish COVID-19 strategy in a random sample of urban older adults. Further strengths include the use of an *ad hoc* questionnaire developed by a multidisciplinary team of experts, and the study sample coming from a well-characterized population-based study. Being based on data from Sweden, the study also provides a unique opportunity for comparison with other settings, as the Swedish strategy against COVID-19 differed from most countries. Certain limitations also need to be highlighted. We did not have recent pre-pandemic measures of physical and mental health. Thus, despite participants being asked to answer the questions for the period since March, the cross-sectional design does not allow us to assess temporal relationship between the onset of the pandemic and the studied outcomes. The findings from this cohort of older adults living in an affluent neighborhood of Stockholm may not be generalizable to the entire Swedish population. However, these findings could be viewed as a best-case scenario, and the identified collateral damage would be expected to be of higher magnitude in less urban and affluent parts of the country.

## Conclusion

The results from this study indicate that, in addition to morbidity and mortality, COVID-19 and its related restrictions during the first half of 2020 have also resulted in changes that negatively affect the health and lives of the elderly living in Central Stockholm. Furthermore, we found age-, sex-, living arrangement- and, to a much lesser extent, education-related differences in the susceptibility to these consequences, with women being at a particularly increased risk. When introducing restrictions, we emphasize the need of a predefined, evidence-based strategy to provide support to those who are most susceptible to these consequences.

## Data Availability

Data are from the SNAC-K project, a population-based study on aging and dementia (http://www.snac-k.se/). Access to these original data is available to the research community upon approval by the SNAC-K data management and maintenance committee. Applications for accessing these data can be submitted to Maria Wahlberg at the Aging Research Center, Karolinska Institutet.

## Contributors

GB, LF, and AC-L developed the study concept and design. GB performed the data analysis and drafted the manuscript. All authors interpreted the data, provided critical revisions and approved the final version of the manuscript for submission.

## Patient consent for publication

Not required.

## Provenance and peer review

Not commissioned; externally peer reviewed.

