## Supplementary Table 1, Supplementary Table 2 for "COVID-19 collateral damage: psychological distress and behavioral changes among older adults during the first outbreak in Stockholm, Sweden"

**Supplementary Table 1. Psychological distress and behavioral changes in the study sample (N=1231) by age and sex during the first COVID-19 outbreak in Stockholm (March-June 2020).**

|  | **Age** | | *p-value* | **Sex** | | *p-value* |
| --- | --- | --- | --- | --- | --- | --- |
|  | Under 80 | Over 80 |  | Male | Female |  |
|  | n=809 | n=422 |  | n=439 | n=792 |  |
| **Psychological distress** | | | | | | |
| **Worried about getting COVID-19** |  |  | 0.003 |  |  | 0.13 |
| Not at all, slightly, moderately | 522 (82.6%) | 510 (88.5%) |  | 378 (87.5%) | 654 (84.3%) |  |
| Very, extremely | 110 (17.4%) | 66 (11.5%) |  | 54 (12.5%) | 122 (15.7%) |  |
| **Worried about loved one getting COVID-19** |  |  | 0.054 |  |  | 0.063 |
| Not at all, slightly, moderately | 462 (72.9%) | 445 (77.7%) |  | 338 (78.2%) | 569 (73.4%) |  |
| Very, extremely | 172 (27.1%) | 128 (22.3%) |  | 94 (21.8%) | 206 (26.6%) |  |
| **Felt nervous/stressed** |  |  | 0.45 |  |  | 0.002 |
| Never, sometimes | 580 (91.3%) | 532 (92.5%) |  | 410 (95.1%) | 702 (90.1%) |  |
| Often or very often | 55 (8.7%) | 43 (7.5%) |  | 21 (4.9%) | 77 (9.9%) |  |
| **Felt lonely^a^** |  |  | 0.19 |  |  | <0.001 |
| To a low extent (<5) | 429 (68.3%) | 361 (64.7%) |  | 319 (74.7%) | 471 (62.1%) |  |
| To a high extent (≥5) | 199 (31.7%) | 197 (35.3%) |  | 108 (25.3%) | 288 (37.9%) |  |
| **Affected by at least one item** | 318 (49.7%) | 290 (49.8%) | 0.96 | 171 (39.1%) | 437 (55.7%) | <0.001 |
| **Reductions in social and physical activities** |  |  |  |  |  |  |
| **Social isolation^b^** | 94 (14.9%) | 101 (17.7%) | 0.19 | 97 (22.8%) | 255 (33.0%) | <0.001 |
| **Reduced light physical activity** | 145 (23.2%) | 207 (36.2%) | <0.001 | 110 (26.0%) | 216 (28.1%) | 0.45 |
| **Reduced vigorous physical activity** | 209 (33.6%) | 117 (20.5%) | <0.001 | 220 (50.5%) | 456 (58.0%) | 0.011 |
| **Affected by at least one item** | 341 (53.3%) | 335 (57.6%) | 0.13 | 83 (19.4%) | 112 (14.5%) | 0.025 |
| **Reductions in care use** | | | | | | |
| **Refrained from seeking medical care** | 66 (10.3%) | 60 (10.3%) | 0.99 | 43 (9.9%) | 83 (10.5%) | 0.71 |
| **Received less home care^c,d^** | 4 (22.2%) | 12 (7.4%) | 0.036 | 2 (4.1%) | 14 (10.7%) | 0.17 |
| **Affected by at least one item** | 69 (10.8%) | 70 (11.9%) | 0.52 | 45 (10.3%) | 94 (11.9%) | 0.40 |
| **Sum of collateral damage dimensions** |  |  |  |  |  |  |
| **0** | 147 (22.9%) | 126 (21.4%) | 0.40 | 123 (28.1%) | 150 (19.0%) | <0.001 |
| **1** | 287 (44.8%) | 250 (42.5%) |  | 207 (47.3%) | 330 (41.7%) |  |
| **2** | 180 (28.1%) | 191 (32.5%) |  | 95 (21.7%) | 276 (34.9%) |  |
| **3** | 27 (4.2%) | 21 (3.6%) |  | 13 (3.0%) | 35 (4.4%) |  |

^a^ Based on UCLA Three-Item Loneliness Scale (range: 3-9)

^b^ Reduction in physical communication without an increase in phone and/or video communication

^c^ Reduction in formal care without an increase in informal care

^d^ Subsample of those who received home care before the pandemic (n=180)

**Supplementary Table 2. Psychological distress and behavioral changes in the study sample (N=1231) by education and living arrangement during the first COVID-19 outbreak in Stockholm (March-June 2020).**

|  | **Education^a^** | | *p-value* | **Living arrangement** | | *p-value* |
| --- | --- | --- | --- | --- | --- | --- |
|  | Low | High |  | Living alone | Not living alone |  |
|  | n=422 | n=809 |  | n=612 | n=616 |  |
| **Worried about getting COVID-19** |  |  | 0.35 |  |  | 0.46 |
| Not at all, slightly, moderately | 360 (86.7%) | 672 (84.7%) |  | 508 (84.7%) | 523 (86.2%) |  |
| Very, extremely | 55 (13.3%) | 121 (15.3%) |  | 92 (15.3%) | 84 (13.8%) |  |
| **Worried about loved one getting COVID-19** |  |  | 0.66 |  |  | 0.004 |
| Not at all, slightly, moderately | 312 (75.9%) | 595 (74.7%) |  | 433 (71.6%) | 473 (78.7%) |  |
| Very, extremely | 99 (24.1%) | 201 (25.3%) |  | 172 (28.4%) | 128 (21.3%) |  |
| **Felt nervous/stressed** |  |  | 0.62 |  |  | 0.042 |
| Never, sometimes | 381 (91.4%) | 731 (92.2%) |  | 566 (93.6%) | 545 (90.4%) |  |
| Often or very often | 36 (8.6%) | 62 (7.8%) |  | 39 (6.4%) | 58 (9.6%) |  |
| **Felt lonely^b^** |  |  | 0.81 |  |  | <0.001 |
| To a low extent (<5) | 269 (67.1%) | 521 (66.4%) |  | 431 (72.4%) | 358 (60.8%) |  |
| To a high extent (≥5) | 132 (32.9%) | 264 (33.6%) |  | 164 (27.6%) | 231 (39.2%) |  |
| **Affected by at least one item** | 213 (50.7%) | 395 (49.3%) | 0.63 | 280 (46.1%) | 326 (53.3%) | 0.012 |
| **Reductions in social and physical activities** |  |  |  |  |  |  |
| **Social isolation^c^** | 76 (18.6%) | 119 (15.0%) | 0.11 | 95 (15.8%) | 100 (16.7%) | 0.67 |
| **Reduced light physical activity** | 129 (31.2%) | 223 (28.4%) | 0.31 | 156 (26.2%) | 196 (32.7%) | 0.013 |
| **Reduced vigorous physical activity** | 94 (22.9%) | 232 (29.7%) | 0.012 | 183 (31.0%) | 142 (23.7%) | 0.005 |
| **Affected by at least one item** | 239 (57.0%) | 437 (54.4%) | 0.38 | 334 (54.9%) | 341 (55.7%) | 0.78 |
| **Reductions in care use** | | | | | | |
| **Refrained from seeking medical care** | 42 (10.1%) | 84 (10.4%) | 0.85 | 60 (9.9%) | 66 (10.8%) | 0.61 |
| **Received less home care^d,e^** | 12 (14%) | 4 (4%) | 0.025 | 3 (6.7%) | 13 (9.6%) | 0.55 |
| **Affected by at least one item** | 52 (12.4%) | 87 (10.8%) | 0.41 | 63 (10.3%) | 76 (12.4%) | 0.27 |
| **Sum of collateral damage dimensions** |  |  |  |  |  |  |
| **0** | 92 (21.8%) | 181 (22.4%) | 0.38 | 141 (23.1%) | 132 (21.4%) | 0.046 |
| **1** | 173 (41.0%) | 364 (45.1%) |  | 285 (46.6%) | 251 (40.7%) |  |
| **2** | 140 (33.2%) | 231 (28.6%) |  | 163 (26.7%) | 207 (33.6%) |  |
| **3** | 17 (4.0%) | 31 (3.8%) |  | 22 (3.6%) | 26 (4.2%) |  |

^a^ Low = elementary, high = high school, university or above

^b^ Based on UCLA Three-Item Loneliness Scale (range: 3-9)

^c^ Reduction in physical communication without an increase in phone and/or video communication

^d^ Reduction in formal care without an increase in informal care

^e^ Subsample of those who received home care before the pandemic (n=180)
